# Long-term storage has minor effects on biobanked neonatal dried blood spot metabolome

**DOI:** 10.1101/2022.11.14.22276667

**Authors:** Filip Ottosson, Francesco Russo, Anna Abrahamsson, Nadia Sara Jensen MacSween, Julie Courraud, Zaki Krag Nielsen, David M. Hougaard, Arieh S. Cohen, Madeleine Ernst

## Abstract

Over 2.5 million neonatal dried blood spots (DBS) are stored at the Danish National Biobank. These samples offer extraordinary possibilities for metabolomics research, including prediction of disease and understanding of underlying molecular mechanisms of disease development. Nevertheless, Danish neonatal DBS have been little explored in metabolomics studies. One question that remains is the long-term stability of the large number of metabolites typically assessed in untargeted metabolomics over long time periods of storage. Here, we investigate temporal trends of metabolites measured in 200 neonatal DBS collected over a time course of 10 years, using an untargeted LC-MS/MS based metabolomics protocol. We found that a majority (79%) of the metabolome was stable during 10 years of storage at -20°C. However, we found decreasing trends for lipid-related metabolites, such as phosphocholines and acylcarnitines. A few metabolites, including glutathione and methionine, may be strongly influenced by storage, with changes in metabolite levels up to 0.1-0.2 standard deviation units per year. Our findings indicate that untargeted metabolomics of DBS samples, with long-term storage in biobanks, is suitable for retrospective epidemiological studies. We identify metabolites whose stability in DBS should be closely monitored in future studies of DBS samples with long-term storage.

## Introduction

Over 2 million neonatal heel prick samples are stored at the Danish National Biobank at Statens Serum Institut and made accessible to researchers worldwide. These samples derive from the population-wide screening for inborn errors of metabolism (IEM), which in Denmark has been performed since the 1980’s. All of these samples can be linked to Danish registry data, containing diverse health and social records, such as disease incidence, operation procedures, age, gender, education, or ethnicity^1,2^, thus providing extraordinary opportunities for research into early diagnosis and detection of diseases. Metabolomics studies in particular could offer unique opportunities to describe potential metabolic etiologies of various diseases. This could provide clinicians with a retrospective tool to investigate disease onset as well as providing researchers with an opportunity to conduct longitudinal cohort studies. Nevertheless, Danish neonatal DBS have been little explored in metabolomics studies. One question that remains uninvestigated is the long term stability of the large number of metabolites typically assessed in an untargeted metabolomics experiment over long time periods of storage. Blood for neonatal screening is usually drawn from the heel, absorbed onto filter paper, and dried for three hours at ambient temperature before analysis and biobank storage^3^. Compared to traditional whole blood sampling, dried blood spot (DBS) samples are less invasive and require less sample volume. DBS samples also have a distinct matrix paper composition and are made of whole blood, which, in conventional blood sampling, is separated into plasma or serum and blood cells/clot. Therefore, both cellular and extracellular compounds are present in DBS, offering multiple opportunities for clinical practice and research.

Long-term stability of DBS samples has been studied to a lesser extent than the corresponding liquid blood drawings, despite being a crucial aspect in order to ensure valid analytical results. DBS samples stored at ambient temperature are prone to considerable short-term changes^4^. For instance, in a panel of 13 amino acids and carnitine species, storage in ambient temperature for 5-15 years resulted in significant changes in concentrations for all metabolites except valine, ranging from 2-28% per year^5^. In particular, acylcarnitines appear to be sensitive to degradation at ambient temperature ^6^. Conversely, storage below -20°C significantly reduces time-dependent metabolite changes in concentrations in a study comparing the 2-year stability of metabolites in DBS at ambient temperature, -20°C and -80°C^7^. However, only a few studies have studied the stability of the DBS metabolome, as measured using untargeted metabolomics. It has been shown that only a small minority of the metabolome in DBS from rats were unstable during one year storage at -20°C ^8^. On the other hand, during storage times up to five years at -20°C, one study found considerable variation in a majority of 6000 measured metabolite features^9^. Further studies, investigating longer storage times, are needed in order to confirm these findings.

In this study, we used an untargeted LC-MS/MS based metabolomics protocol to identify long term temporal trends in thousands of metabolites in DBS samples stored at the Danish National Biobank at -20°C from one to ten years.

## Material and Methods

### Study design

We retrieved from the Danish National Biobank a cohort of 200 neonatal DBS stored over a time period of ten years at -20°C (2010-2019). 20 neonatal DBS were retrieved for each year (ten females, ten males). To control for variation introduced by other sources than storage time, we selected DBS from children born in July, sampled at two days of age, and born at 40 weeks of gestation at Hvidovre Hospital, Denmark. The study was conducted in accordance with the Declaration of Helsinki, and the protocol complies with the Danish Ethical Committee law by not being a health research project (Section 2,1), but a method development study not requiring an ethical approval^2^. The Committees on Health Research Ethics for the Capital Region of Denmark waived ethical approval for this work.

### Sample Preparation

All samples (including blank, pooled, and external quality control samples) were submitted to untargeted metabolomic profiling using liquid chromatography tandem mass spectrometry (LC-MS/MS) at Statens Serum Institut, Copenhagen, Denmark, in December 2020. For LC-MS/MS data acquisition, samples were randomly distributed over three 96-well plates (batches). A large batch of DBS consisting of adult blood from a single individual was prepared at the beginning of the study, and stored at -20°C. Aliquots (3.2-mm-diameter punches) were distributed on all plates and used as external controls (EC). Each plate consisted of 2 water blanks, 8 EC, 4 paper blanks (PB, 3.2-mm-diameter punches of blank filter paper), 4 pooled samples (equal aliquots of all samples within a plate), and 67 samples for the second and third plate and 66 samples for the first plate. All solvents were LCMS-grade, and were purchased from Thermo Fisher Scientific (Waltham, MA, USA). DBS samples (3.2-mm-diameter punches) were dissolved in 80% methanol in water and 100 µL extracts were placed in 96-well plates. All extraction steps were performed on a Microlab STAR automated liquid handler (Hamilton Bonaduz AG, Bonaduz, Switzerland).

### Metabolomics profiling

The LC-MS/MS platform consisted of a timsTOF Pro mass spectrometer with an Apollo II ion-source for electrospray ionization (Bruker Daltonics, Billerica, MA, US) coupled to a UHPLC Elute LC system (Bruker Daltonics). The chromatographic separation system included a binary pump, an autosampler with cooling function, and a column oven with temperature control. For infusion of the reference solution, used for external and internal mass calibration, an additional isocratic pump, Azura Pump P4.1S (Knauer, Berlin, Germany) was used. The analytical separation was performed on an Acquity HSS T3 (100 Å, 2.1 mm x 100 mm, 1.8 µm) column (Waters, Milford, MA, US). The mobile phase consisted of solvent A (99.8% water and 0.2% formic acid) and B (49.9% methanol, 49.9% acetonitrile and 0.2% formic acid). The analysis started with 99% mobile phase A for 1.5 min, followed by a linear gradient to 95% mobile phase B during 8.5 min, an isocratic flow at 95% mobile phase B for 2.5 min before going back to 99% mobile phase A and equilibration for 2.4 min. Total run time for each injection was 15 min and the analysis time for a full 96-well plate was approximately 25 h. Samples were maintained at +15°C in the autosampler, 5 µL were loaded onto the column with a flow rate of 0.4 mL/min and a column temperature of 40°C.

Tandem mass spectrometric analysis was performed in Q-TOF mode with TIMS off, and auto MS/MS using the following settings: ionization mode set to positive ionization, mass range set to 20–1000 *m/z* and a Spectra Rate of 3 Hz. Source settings as Capillary: 2500 V, Nebulizer Gas: 2.5 Bar, Dry Gas flow: 8 L/min, Dry Gas temperature: 240°C. Tune settings were as follows: Funnel 1 RF and Funnel 2 RF: 200Vpp, isCID: 0 eV, Multipole RF: 60 Vpp, Deflection Delta: 60 V, Quadrupole Ion Energy: 5 eV with a low mass set to 60 *m/z*, Collision Cell Energy set to 7 eV with a pre Pulse Storage of 5 µs. Stepping was used in Basic Mode with a Collison RF from 250 – 750 Vpp, Transfer Time 20 – 50 µs and Timing set to 50% for both. For MS/MS, only the collision energy ranged from 100%to 250% with timing set to 50% for both. Auto MS/MS was used with a predefined Cycle Time of 0.5 s, Active Exclusion was used with Exclusion after 3 Spectra and a Release time set to 0.20 min. Sodium formate clusters were applied for instrument mass calibration and for internal recalibration of individual samples. A Precursor Exclusion list was used with Exclusion of mass range of 20-60 *m/z*.

### Metabolomics preprocessing

Bruker .d files were exported to the .mzML format using ProteoWizard’s MSConvert^10^, and subsequently preprocessed using the Ion Identity Network workflow in MZmine^11,12^ (version 2.37.1.corr17.7).

Data was cropped, with chromatogram retention time from 0.4 to 12 min and *m/z* range from 0 to 1100 retained. Mass lists were then created with MS1 intensity above 5E2 and MS2 intensity above 0 retained. The chromatogram was built through the ADAP chromatogram builder^7^ by using the following parameters: minimum group size of scans: 3, group intensity threshold: 5E2, minimum highest intensity: 1E3, and *m/z* tolerance: 0.002 *m/z* or 5 ppm. The chromatogram was smoothed with a filter width of 5 and further deconvoluted using the MEDIAN *m/z* center calculation with *m/z* range for MS2 scan pairing: 0.002 Da and retention time range for MS2 scan pairing: 0.3 min. The local minimum search algorithm was used for deconvolution with parameters set to: chromatographic threshold: 85%, minimum RT range: 0.01 min, minimum relative height: 0%, minimum absolute height: 1E3, min ratio of peak top/edge: 2.2, peak duration range: 0.01-0.5 min. The peaks were deisotoped using the isotopic peak grouper function, with parameters set to: *m/z* tolerance: 0.002 *m/z* or 5 ppm, retention time tolerance: 0.3 min, monotonic shape: on, maximum charge: 2, representative isotope: most intense. Peaks from all samples were aligned using the join aligner function with parameters set to: *m/z* tolerance: 0.002 *m/z* or 5 ppm, retention time tolerance: 0.5 min, weight for *m/z*: 75%, weight for retention time: 25%. Rows were then filtered using the duplicate peak filter with the new average filter mode and *m/z* tolerance set to 0.001 *m/z* or 5 ppm and RT tolerance to 0.03 min. The metaCorrelate function was used to find correlating peak shapes with parameters set to: RT tolerance: 0.1 min, min height: 1E3, noise level: 5E2, min samples in all: 2 (abs), min samples in group: 0 (abs), min %-intensity overlap: 60%, exclude estimated features (gap-filled): on. Parameters for the correlation grouping were set as follows: min data points: 5, min data points on edge: 2, measure: Pearson, min feature shape correlation: 85%. Ion identity networking parameters were set to: *m/z* tolerance: 0.002 *m/z* or 5 ppm, check: one feature, min height: 1E3 with ion identity library parameters set to: MS mode: positive, maximum charge: 2, maximum molecules/cluster: 2, adducts: M+H, M+Na, M+K, modifications: M-H2O, M-NH3. Further ion identity networks were added with *m/z* tolerance: 0.002 *m/z* or 5 ppm, min height: 1E3 and ion identity library parameters set to: MS mode: positive, maximum charge: 2, maximum molecules/cluster: 6, adducts: M+H, M+Na, modifications: M-H2O, M-2H2O, M-3H2O, M-4H2O, M-5H2O and *m/z* tolerance: 0.002 *m/z* or 5 ppm, min height: 1E3, and annotation refinement on with parameters set to: delete smaller networks: link threshold: 4, delete networks without monomer: on, and ion identity library parameters set to MS mode: positive, maximum charge: 2, maximum molecules/cluster: 2, adducts: M+H, M+Na, M+K, modifications: M-H2O, M-NH3. Finally, two feature tables were exported in the .csv format: one feature table containing all extracted mass spectral features and another feature table filtered for mass spectral features with associated fragmentation spectra (MS2). An aggregated list of MS2 fragmentation spectra was exported in the .mgf format and submitted to ion identity feature-based mass spectral molecular networking through the Global Natural Products Social Molecular Networking Platform (GNPS)^12–14^.

Before statistical analysis, mass spectral features with a relative intensity less than 20 times the mean relative intensity of all paper blank samples were removed. Relative intensities were further batch normalized through centering by subtracting the column means (omitting NAs) of each batch and scaling by the standard deviation. Missing values were thereafter imputed using the OptSpace matrix completion algorithm implemented in the robust Aitchison open-source software DEICODE, implemented in Qiime2^15^, assuming a rank of 100 for the underlying low-rank structure^16^. Furthermore we removed features present in less than 5% of the samples, resulting in a final cohort of 200 samples and 731 metabolic features.

### Metabolite identification

To annotate mass spectral features to putative chemical structures, a mass spectral molecular network was created through the GNPS Platform (http://gnps.ucsd.edu) using the ion identity feature based molecular networking workflow (https://ccms-ucsd.github.io/GNPSDocumentation/fbmn-iin/)^12–14^. The data was filtered by removing all MS/MS fragment ions within +/-17 Da of the precursor *m/z*. MS/MS spectra were window filtered by choosing only the top 6 fragment ions in the +/-50 Da window throughout the spectrum. The precursor ion mass tolerance was set to 0.02 Da and a MS/MS fragment ion tolerance of 0.02 Da. A network was then created where edges were filtered to have a cosine score above 0.7 and more than 4 matched peaks. Further, edges between two nodes were kept in the network if and only if each of the nodes appeared in each other’s respective top 10 most similar nodes. Finally, the maximum size of a molecular family was set to 100, and the lowest scoring edges were removed from molecular families until the molecular family size was below this threshold. The spectra in the network were then searched against all GNPS’ spectral libraries. The library spectra were filtered in the same manner as the input data. All matches kept between network spectra and library spectra were required to have a score above 0.7 and at least four matched peaks.

To further enhance chemical structural information within the molecular network, substructure information was incorporated into the network using the GNPS MS2LDA workflow (https://ccms-ucsd.github.io/GNPSDocumentation/ms2lda/)^17–19^. Furthermore, information from *in silico* structure annotations from Network Annotation Propagation^20^ and Sirius+CSI:FingerID^21^ were incorporated into the network using the GNPS MolNetEnhancer workflow (https://ccms-ucsd.github.io/GNPSDocumentation/molnetenhancer/)^22^. Chemical class annotations were performed using deep neural networks in CANOPUS^23^ and followed the ClassyFire chemical ontology^24^.

### Statistical Analysis

The overall variation in the metabolite data was analyzed using principal component analysis (PCA), in R package *mixOmics*^*25*^. To assess associations between principal components and storage time, we performed linear regression models. To identify metabolic features significantly increasing or decreasing over time, we performed a linear regression analysis for each metabolite individually. All linear regression models were adjusted for sex, birth weight and the mothers’ age. P-values were adjusted for multiple hypothesis testing using the false discovery rate (FDR) method^26^. To visualize temporal trends, we calculated the median for all significant metabolites (linear regression, FDR-adjusted P-value < 0.05) and subtracted the most recent sampling year (2019), which was used as baseline. Results from linear regression models were confirmed using non-parametric correlation tests using Spearman’s ρ. Also, longitudinal trends in metabolite data were explored by multivariate statistics using the R package *timeOmics*^*27*^. Briefly, longitudinal changes in metabolite levels were modeled using linear mixed model splines. By applying PCA on modeled metabolite profiles, metabolites with similar longitudinal trends could be clustered together. The optimal number of principal components were optimized by maximizing the silhouette coefficient. All statistical analyses were performed in R 4.1.1 or Python 3.7. Code and Jupyter notebooks are publicly accessible at: https://github.com/SSI-Metabolomics/Temporal_SupplementaryMaterial/.

### Putative identification of degradation products

To identify putative degradation products we performed a pair-wise correlation analysis of all metabolic features correlating significantly with year of sampling (FDR-adjusted P-value < 0.05) using Pearson’s ρ. Putative degradation was then defined as two metabolic features, which 1) correlate significantly with year of sampling (FDR-adjusted P-value < 0.05), 2) correlate negatively with each other (Pearson’s r < 0; P-value < 0.05), 3) exhibit chemical structural relationship either through high tandem mass spectral similarity (cosine > 0.7) or shared MS2LDA substructural motifs. For visualization and chemical structural annotation, putative degradation products were identified within the mass spectral molecular network by adding edges (connecting lines) between two nodes, meeting criteria 1), 2) and 3).

## Results

A total of 731 metabolites (mass spectral features with unique MS/MS fragmentation patterns) were measured and present in at least 5% of the samples. Putative annotation on the metabolite class level was conducted by combining mass spectral molecular networking (GNPS), unsupervised substructure discovery (MS2LDA), *in silico* annotation through Network Annotation Propagation^20^ Sirius+CSI:FingerID, MolNetEnhancer, and deep neural network in CANOPUS. This resulted in chemical class annotation (level 3 annotation^29^) for 182 metabolites (24.9%).

To examine whether overall variation in the metabolomics data was related to storage time of DBS samples, PCA was performed. The first principal component (PC) explained 11.5 % of the variation in the data set and the cumulative explained variance of the first four PCs was 29.2% (Figure S1A). Samples with similar storage time did not cluster according to the PCs (Figure S1B), but PC2 (beta=0.068, p=5.5e-3), PC3 (beta=0.090, p=2.1e-4) and PC4 (beta=0.12, p=1.6e-7) were significantly (p<0.05) associated with storage time in linear regression models adjusted for sex, birth weight and age of the mother (Figure S1C).

We proceeded to explore the association between each individual metabolite and storage time. In linear regression models, adjusted for sex, birth weight and age of the mother, 152 out of 731 metabolites (20.8%) were significantly (FDR-adjusted P-value<0.05) associated with storage time (Figure 1A) (Table S1). Out of these, 71 were inversely associated with storage time and 81 showed a positive association. Comparing the metabolite levels after ten years of storage with one year of storage, the median metabolite level was on average 0.67 standard deviations higher for metabolites with significant positive beta coefficients and 0.64 standard deviations lower for metabolites with significant negative beta coefficients (Figure 1B). In total, 46 of the 152 significant metabolites (30%) could be assigned to a metabolite class (Figure 1C) (level 3).

**Figure 1.**
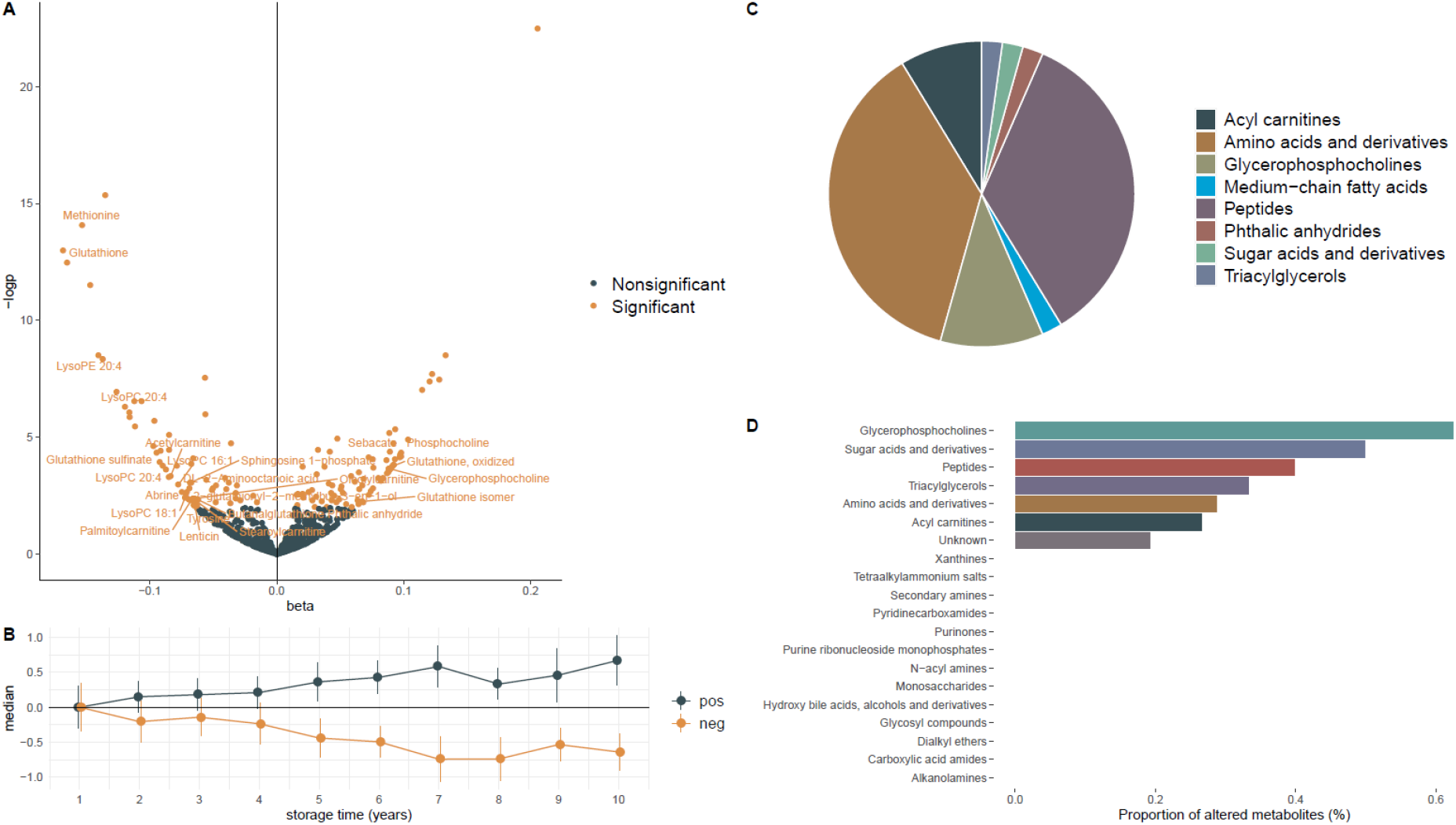
Beta coefficients from linear regression models **(A)**, indicating change for each metabolite in standard deviation units per year of storage time. Significant associations indicate false discovery-rate P-value <0.05. **(B)** Median levels of all the 152 metabolites significantly associated with storage time for each year of storage time. Point estimates show mean value at each time point and direction (pos: positive, neg: negative), and error-bars indicate standard deviations. **(C)** Metabolite classes for annotated metabolites significantly associated with storage time. **(D)** Proportion of metabolites within each class associated with storage time, out of 20 putatively annotated classes, only 7 showed significant alterations with storage time.

Among the eight represented metabolite classes, four contributed with more than one metabolite, including amino acids and derivatives (N=17), peptides (N=16), glycerophosphocholines (N=5) and acylcarnitines (N=4). In total, 24 significant metabolites achieved at least a second level annotation^29^, among which the strongest positive association with storage time was seen for glycerophosphocholine (beta=0.098, p=4.7e-5) and the strongest negative association for methionine (beta=-0.15, p=6.3e-12) and glutathione (beta=-0.18, p=7.7e-11). Overall, 13 classes (with at least two measured metabolites) had no metabolites significantly associated with storage. Among classes with metabolites associated with storage, the proportions of associated metabolites ranged from 27 % (acyl carnitines) to 63 % (glycerophosphocholines). For metabolites without class annotation, 19 % were associated with storage time (Figure 1D).

Metabolites annotated as peptides or amino acids and derivatives did not show similar temporal patterns within each respective class. All acylcarnitines showed negative associations with storage. For glycerophosphocholines, all lysophosphatidylcholine species were negatively associated with storage, while levels of the head group glycerophosphocholine were positively associated with storage (Figure 2).

**Figure 2.**
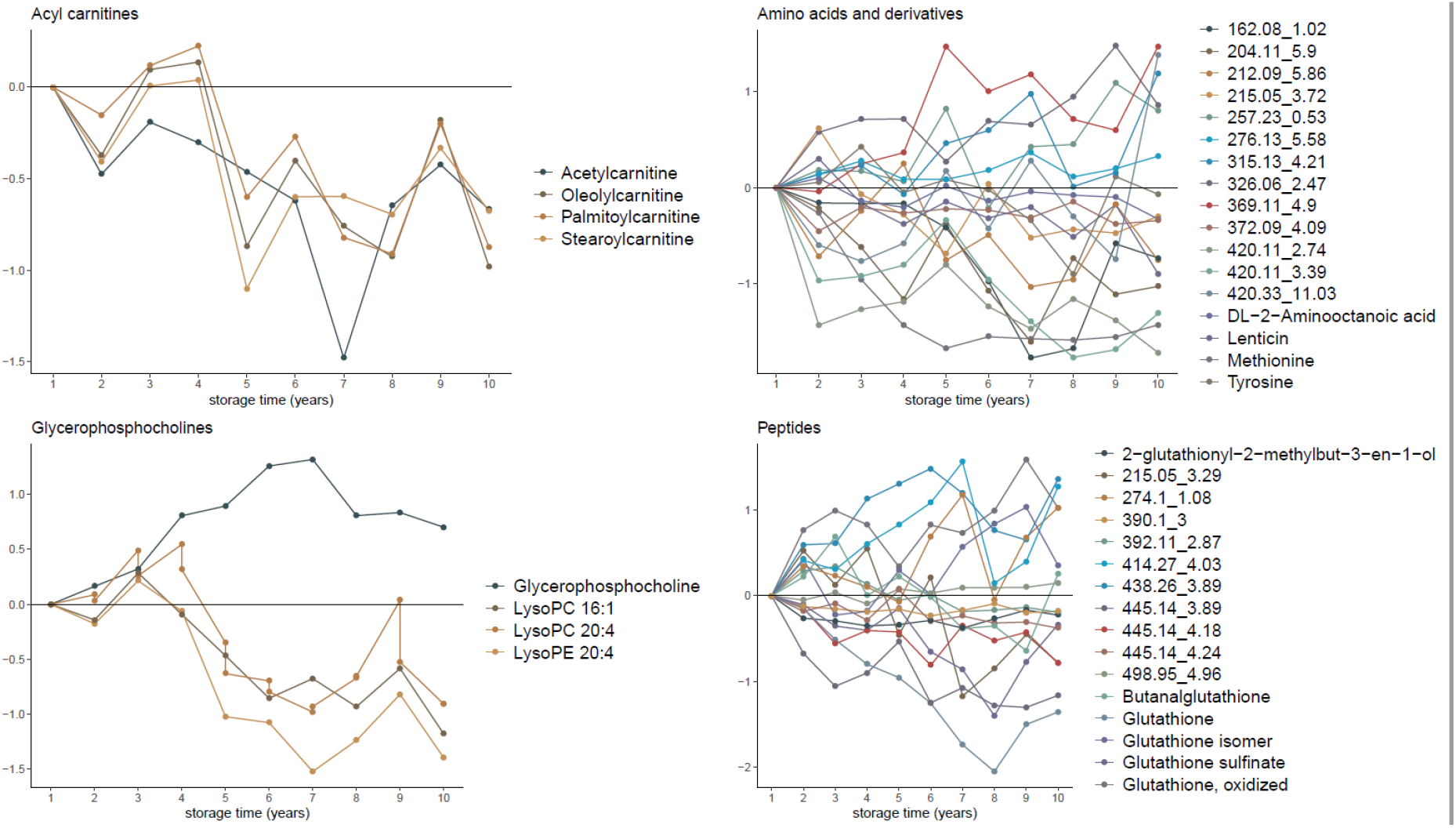
Median metabolite levels for different storage times. Data are shown for all four metabolite classes with more than one metabolite significantly associated with storage time. Metabolites without level 2 annotations are labeled according to their *m/z* and retention time (mz_rt).

**Figure 3.**
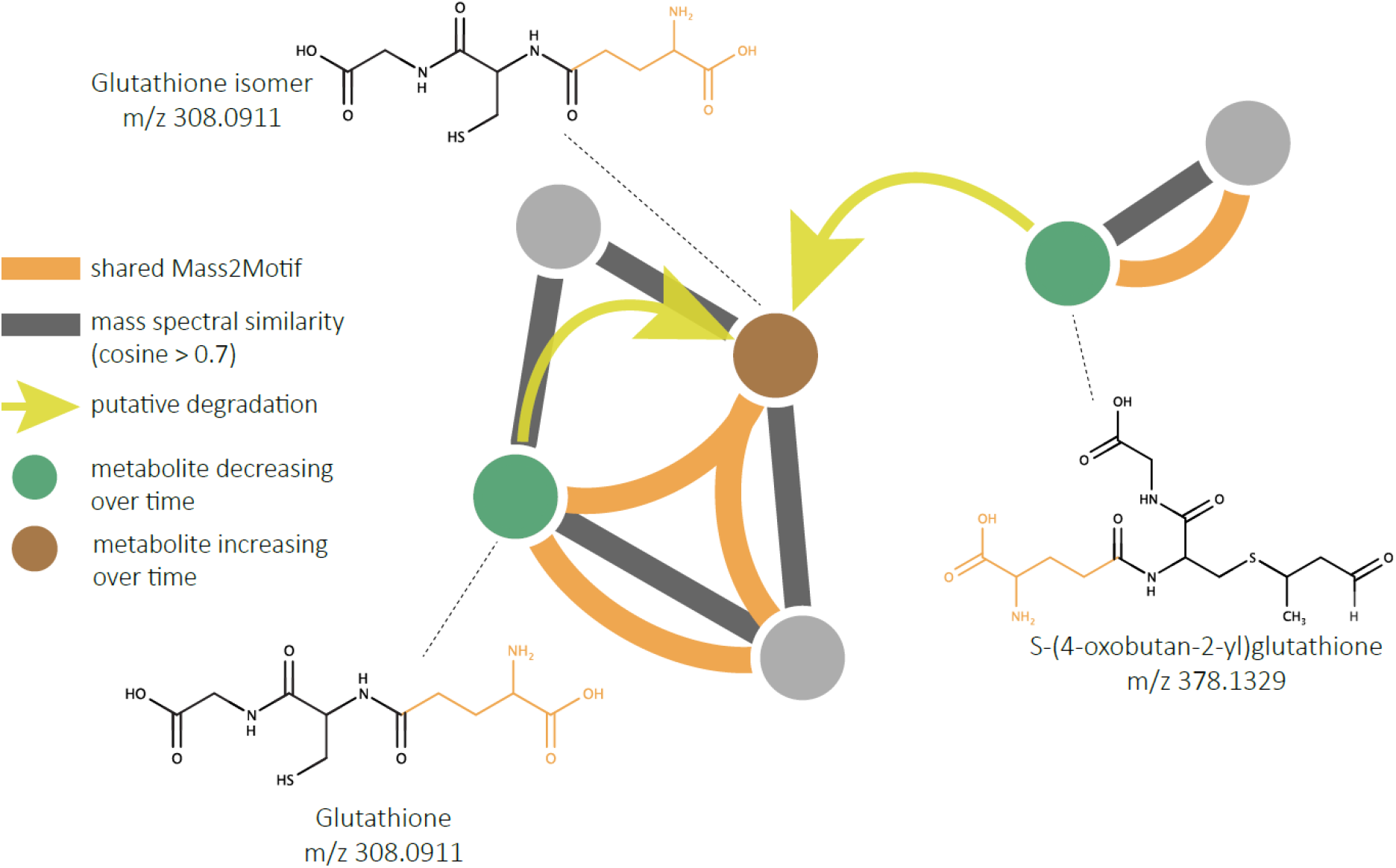
Putative degradation of glutathione structural analogues. Putative degradation was identified through the mass spectral molecular network and defined as two metabolic features, which 1) correlate significantly with year of sampling (FDR-adjusted P-value < 0.05), 2) correlate negatively with each other (Pearson’s r < 0; P-value < 0.05), 3) exhibit chemical structural relationship either through high tandem mass spectral similarity (cosine > 0.7) or shared MS2LDA substructural motifs. Shared MS2LDA substructural motifs are further indicated in orange in the molecular structures.

Overall, similar results were achieved when comparing the results from the linear regression models with those from Spearman’s correlation coefficient tests, where 141 metabolites were significantly (FDR-adjusted P-value<0.05) correlated with storage time, out of which 128 were significant using linear regression (Figure S2).

We next explored metabolite changes upon storage using a multivariate approach implemented in the *timeOmics* R package^27^. The optimal clustering was achieved when using only the first principal component (PC1). PC1 was strongly associated with storage time, explaining 81 % of the variation in storage time. The loadings of PC1 were very similar to the beta-coefficients of the linear regression models (Pearson’s ρ = 0.93). For instance, glutathione was the metabolite with the strongest (negative) contribution to PC1 and among the metabolites with strongest association in the linear regression models (Figure S3).

Observing both positive and negative temporal trends for metabolites, we sought to identify putative transformation pathways for metabolites in our dataset. We found one, and in this putative pathway, an unknown glutathione isomer seems to be formed either by deconjugation of crotonaldehyde from S-(4-oxobutan-2-yl)glutathione or directly from glutathione. This is supported by temporal trends, mass spectral similarities and shared Mass2Motiff. Glutathione (beta=-0.17, P=1E-13) and S-(4-oxobutan-2-yl)glutathione (beta=-0.06, P=4.8e-3) are decreasing in levels with longer storage time, while levels of the unknown glutathione isomer are increased with longer storage time (beta=0.09, P=1.8E-5).

## Discussion

In order to study the long-term stability of untargeted metabolomics data in DBS stored at -20°C, we investigated the relation between over 700 metabolite features and sample storage up to ten years. A minority of the metabolites (20.8%) were significantly associated with storage time, indicating that DBS samples stored in biobanks over an extensive time at -20°C are suitable for untargeted metabolomics studies. The levels of a few metabolites, such as glutathione and methionine, may be heavily influenced by extended storage time in DBS and should be closely monitored in future studies.

Applying untargeted metabolomics in prospective or retrospective cohorts has large potential, but also several challenges, including effects of long-term storage times. The plasma metabolome has been seen to be stable up to five years of storage at -80°C^30^, given that samples do not go through several freeze-thaw cycles^31^. Recently, it was shown that among 200 plasma metabolites, only 2% were significantly altered after seven years, but 26% upon 16 years of storage at -80°C^32^.

As opposed to plasma samples, DBS are usually stored at -20°C or even at room temperature, emphasizing the need to ensure the stability of long-term storage. A previous study by Li and collaborators has shown that 76% of DBS metabolites in a targeted metabolomics panel were influenced by storage in -80°C up to one year. The major effect of storage was seen between months one and three, where influenced metabolites on average decreased in concentration to 60% of the original concentration, while small concentration changes were seen between months three and twelve^33^. There are few studies of metabolite stability in DBS over a longer time span than one year. In a targeted metabolomics study, with several samples collected over two years, alterations in metabolite stability could only be seen in samples stored at room temperature, as opposed to -20°C^7^. Rus and collaborators applied untargeted metabolomics in six individuals, who deposited DBS samples each year over a period of six years. The authors showed that 30-35% of the ∼6,000 measured metabolite features had a between-sample coefficient of variation (CV) < 20%, while the majority of metabolite features displayed large alterations over time^9^.

In order to best capture the effect of storage, we chose to evaluate the temporal trend in the present study using linear regression models, assuming linear changes in metabolite levels over time. As opposed to the previous study by Rus and collaborators, our investigation showed that almost 80% of the measured metabolite features were unrelated to storage time. Among the 152 metabolites that were significantly associated with storage time, we observed that the levels of around half of the metabolites increased with storage, while the remaining decreased. This is in contrast to what was previously seen during one year of storage, where a large majority of influenced metabolites in a targeted panel decreased with time^33^. Our finding that around half of the metabolites increase with storage and half decrease indicates that chemical transformations may occur over time.

To investigate chemical transformations we combined mass spectral molecular networking, *in silico* structure and substructure annotation with information on metabolites significantly increasing or decreasing over time. We found a potential transformation pathway involving glutathione, a compound that is previously known to be prone to degradation in DBS samples^33^. The transformation involves formation of an unidentified glutathione isomer, from glutathione degradation and/or by deconjugation of S-(4-oxobutan-2-yl)glutathione. Moreover, glutathione may also be oxidized to form a disulfide dimer, a process which is well-known in nature, where glutathione is a potent scavenger of reactive oxygen species^34^. Our findings indicate that oxidation could contribute to glutathione degradation, since oxidized glutathione is increasing with storage time. On the other hand, glutathione and oxidized glutathione were not inversely correlated with each other.

Although a large majority of the metabolites in our study were not significantly associated with storage, relatively strong associations were seen for some metabolites, such as glutathione and methionine. For instance, our results indicate that the levels of glutathione decrease at a rate of approximately 0.1-0.2 standard deviation units per year, resulting in significant imprecision when analyzing samples with very different storage time. Overall, our results are consistent with previous studies where several of the metabolites, which are associated with storage time in the present study, have previously been reported to be unstable in DBS samples, including glutathione^33^, Sphingosine-1-phosphate^33^ and palmitoylcarnitine^33^.

Additionally we observed that acylcarnitines and lysophosphocholines were inversely associated with storage time, indicating either residual enzymatic degradation of the fatty-acids or non-enzymatic hydrolysis of the head-groups upon extended storage. Consistent with the latter, we observed that the head group of lysophosphocholines, glycerphosphocholine, was positively associated with storage and a similar trend was seen for free carnitine, although not statistically significant after multiple-test correction. Previous studies display conflicting findings regarding storage of lysoPCs, where two studies in plasma have shown either increasing^32^ and decreasing^30^ concentrations of lysoPCs upon storage. In DBS, several lysoPCs have been shown to decrease in concentration during one year of storage^33^, but some lysoPC species, such as LysoPC C26:0, appear to be stable^35^. In our study glycerophosphocholines was the metabolites class with the highest proportion (63 %) of metabolites associated with storage time. Decreasing concentrations of acylcarnitines has been reported in several studies of both plasma^30^ and DBS^33^ samples, but no evidence of increases in free carnitine has been found previously. It is noteworthy that although we see a negative temporal trend for acylcarnitines in this study, the proportion of altered acyl carnitines (27 %) was close to the overall average (21 %), indicating that the stability may not be worse for acyl carnitines than for other metabolite classes.

In the present study, we aimed at separating the temporal effects of storage from biological variation by analyzing neonatal DBS samples, with identical gestational age, age of sampling and birth month. Moreover, in the regression analysis we adjusted for other potential confounding factors, such as birth weight, sex and the age of the mother. Despite these efforts, a residual biological variation may result in statistically significant differences between individual time points that are not related to storage per se. To minimize the risk of false positives, we modeled a linear association between metabolite and storage, disregarding differences between two individual time points. Results from the linear regression models were confirmed using alternative approaches using both Spearman’s correlation tests and multivariate statistics.

When using data from metabolites that are prone to degradation in epidemiological studies, it may be preferable to adjust for storage time in statistical models. In that light, this study can serve as a reference for identifying metabolites whose levels may be in need of adjustments when comparing samples with very different storage times.

## Conclusions

A large majority of the DBS metabolome is stable during storage at -20°C for up to ten years. A few metabolites, including methionine and glutathione, may be strongly influenced by storage, and should be closely monitored in metabolomics studies of DBS. Overall, our findings indicate that untargeted metabolomics can be applied in retrospective studies of DBS samples with long-term storage in biobanks.

## Data Availability

Data generated and/or analyzed in this study are not publicly available due to the risk of compromising individual privacy but are available from the corresponding author on reasonable request and provided that an appropriate collaboration agreement can be agreed upon.

## ASSOCIATED CONTENT

### Supporting Information

### Author information

## Author Contributions

F.O. performed statistical analysis and manuscript drafting, contributed to chemical structural annotation, study design and data interpretation. F.R. contributed to the study conceptualization, design, data interpretation, statistical analysis and manuscript draft. M.E. preprocessed LC-MS/MS data, performed statistical analysis and chemical structural annotation, created transformation networks and contributed to study conceptualization, design, data interpretation and manuscript draft. A.A. and N.S.J. acquired LC-MS/MS data and contributed to the preprocessing. Z.K.N and J.C. contributed to the study design, conceptualization and data acquisition. A.C. and D.H. conceptualized and designed the study, obtained funding and take full responsibility for compliance with data sharing policies and ethical approval of the study. All authors critically revised the manuscript for important intellectual content.

## Acknowledgements

This research has been conducted using the Danish National Biobank resource supported by the Novo Nordisk Foundation. We thank Susan Svane Laursen for technical assistance and Cameron Martino and James T. Morton for support in the usage and assessment of DEICODE.

## Abbreviations

LC: liquid chromatography
MS: mass spectrometry
MS/MS: tandem mass spectrometry
TIMS: trapped ion mobility spectrometry
TOF: time-of-flight
Q-TOF: quadrupole time-of-flight.

---

**Figure S1.**
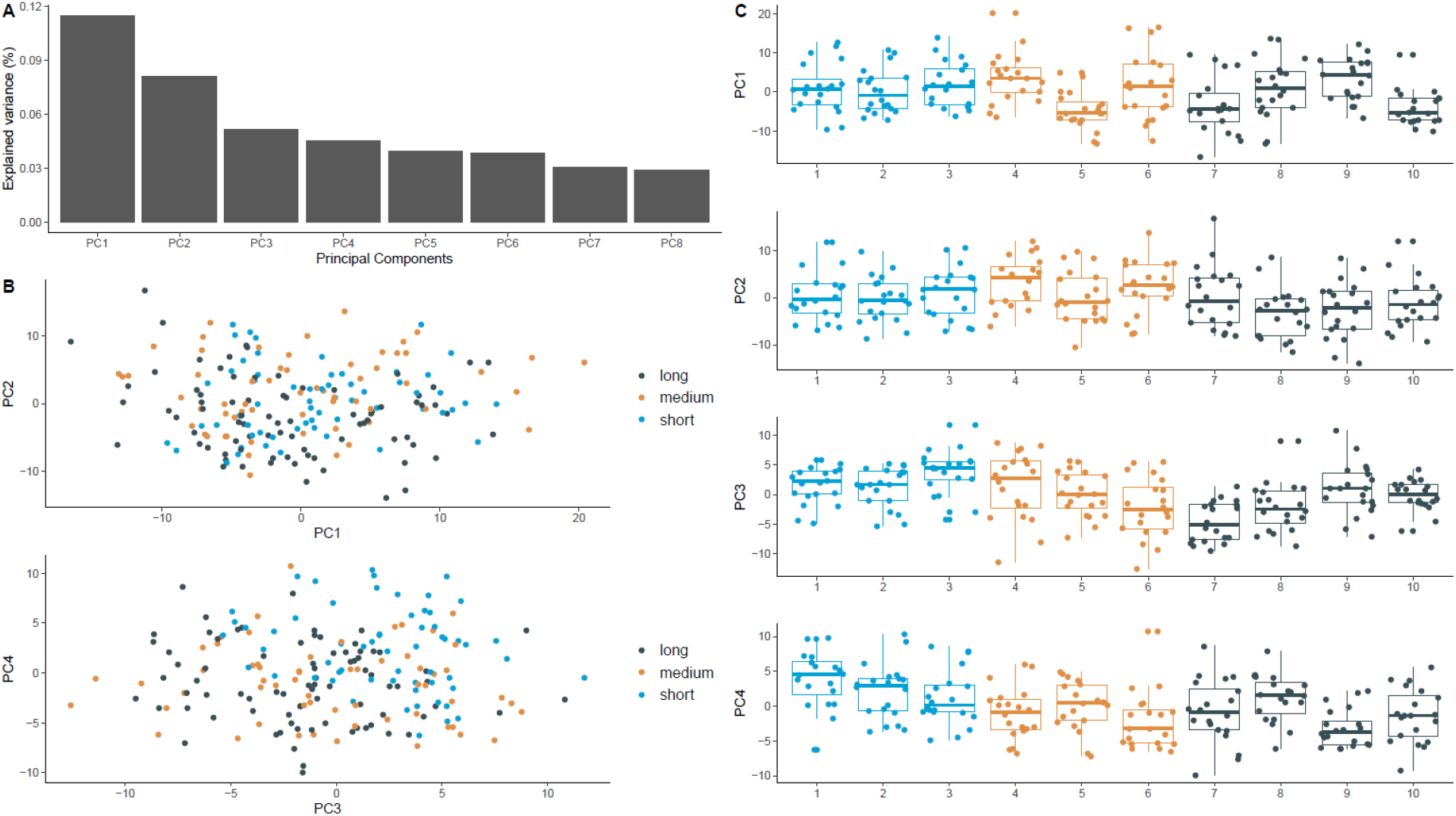
Principal component analysis of metabolite data from DBS of 200 neonates, showing the explained variance for the first 8 principal components (PC) **(A)**. Values for PC1-PC4 for short storage time (1-3 years), medium storage time (4-6 years) and long storage time (7-10 years) **(B)**. Boxplots indicate median, 25^th^ and 75^th^ percentiles for each PC and storage time **(C)**.

**Figure S2.**
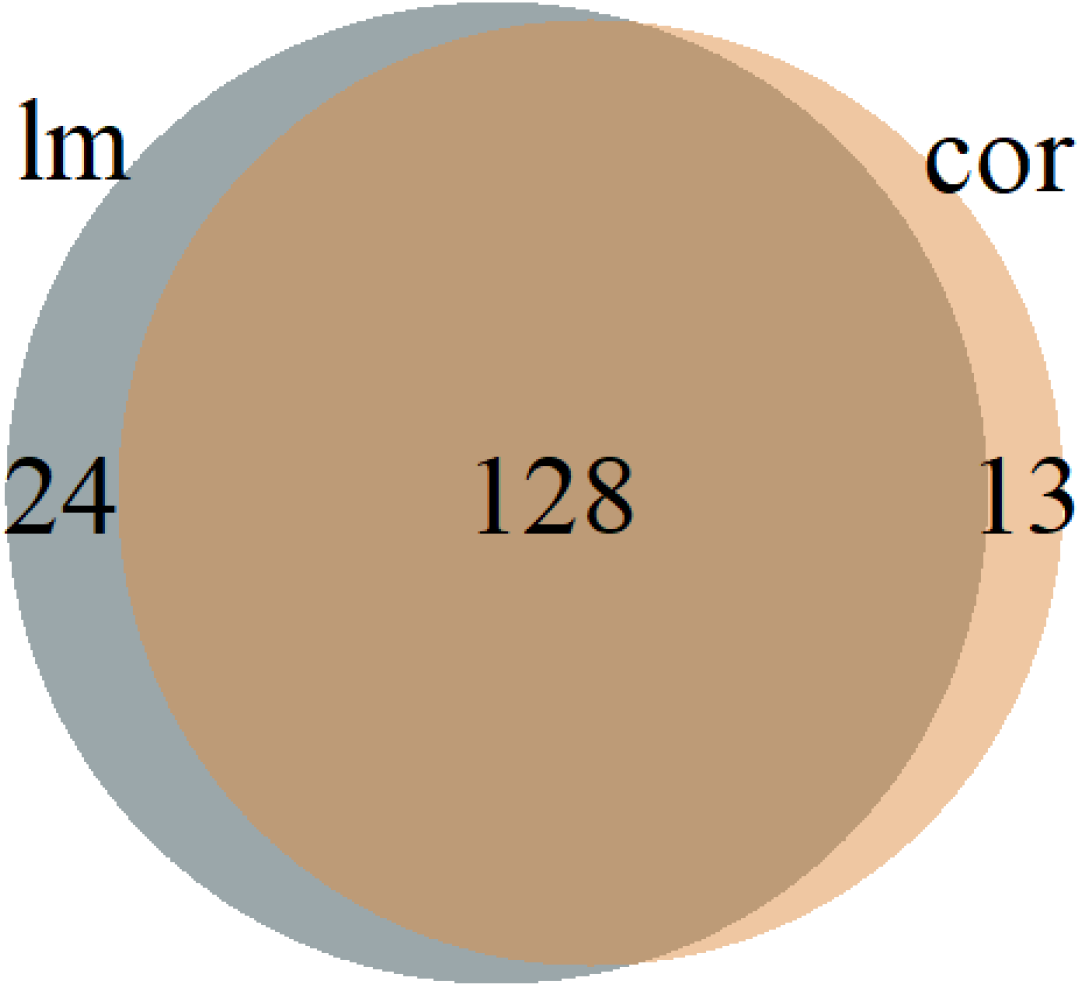
Venn Diagram of number of metabolites significantly (FDR-adjusted P-value<0.05) associated with storage time, using either linear regression models adjusted for sex, birth weight and age of the mother (lm) or Spearman’s correlation test (cor).

**Figure S3.**
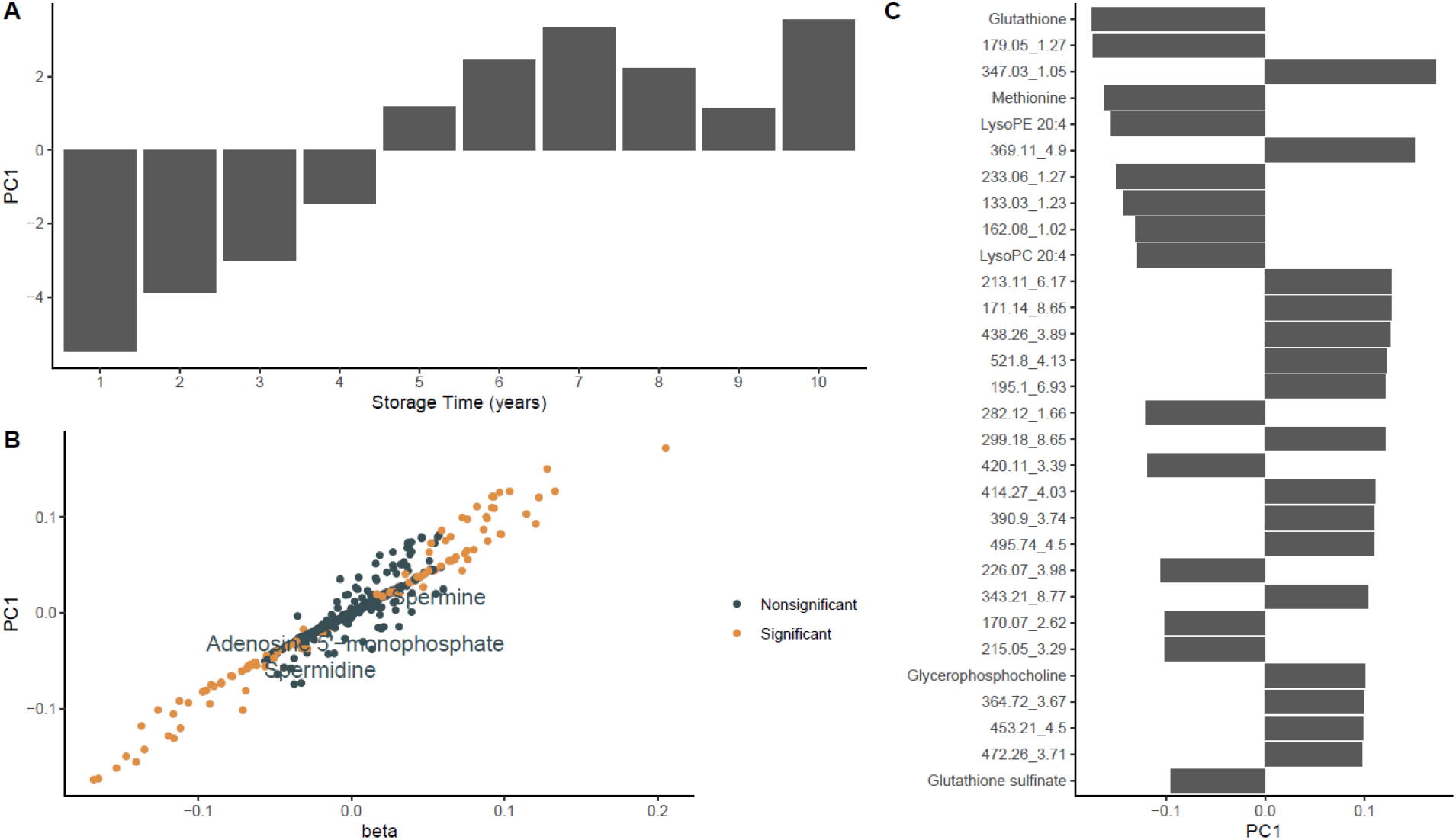
Multivariate modeling of temporal trends found strong association between a principal component of longitudinal changes in metabolite levels (PC1) and storage time **(A)**. The loadings of PC1 were similar to beta-coefficients from linear regression models on storage time. Individual metabolites are labeled according to significance in the linear regression analyses **(B)**. The loadings of PC1 indicated that glutathione was the metabolite with the strongest contribution to the clustering **(C)**.

## References

1. Thygesen, L. C., Daasnes, C., Thaulow, I. & Brønnum-Hansen, H. Introduction to Danish (nationwide) registers on health and social issues: structure, access, legislation, and archiving. Scand. J. Public Health 39, 12–16 (2011).

2. Nørgaard-Pedersen, B. & Hougaard, D. M. Storage policies and use of the Danish Newborn Screening Biobank. J. Inherit. Metab. Dis. 30, 530–536 (2007).

3. Wilhelm, A. J., den Burger, J. C. G. & Swart, E. L. Therapeutic drug monitoring by dried blood spot: progress to date and future directions. Clin. Pharmacokinet. 53, 961–973 (2014).

4. Michopoulos, F., Theodoridis, G., Smith, C. J. & Wilson, I. D. Metabolite profiles from dried blood spots for metabonomic studies using UPLC combined with orthogonal acceleration ToF-MS: effects of different papers and sample storage stability. Bioanalysis 3, 2757–2767 (2011).

5. Strnadová, K. A. et al. Long-term stability of amino acids and acylcarnitines in dried blood spots. Clin. Chem. 53, 717–722 (2007).

6. Fingerhut, R. et al. Stability of acylcarnitines and free carnitine in dried blood samples: implications for retrospective diagnosis of inborn errors of metabolism and neonatal screening for carnitine transporter deficiency. Anal. Chem. 81, 3571–3575 (2009).

7. Prentice, P., Turner, C., Wong, M. C. & Dalton, R. N. Stability of metabolites in dried blood spots stored at different temperatures over a 2-year period. Bioanalysis 5, 1507–1514 (2013).

8. Palmer, E. A., Cooper, H. J. & Dunn, W. B. Investigation of the 12-Month Stability of Dried Blood and Urine Spots Applying Untargeted UHPLC-MS Metabolomic Assays. Anal. Chem. 91, 14306–14313 (2019).

9. Rus, C.-M., Di Bucchianico, S., Cozma, C., Zimmermann, R. & Bauer, P. Dried Blood Spot (DBS) Methodology Study for Biomarker Discovery in Lysosomal Storage Disease (LSD). Metabolites 11, (2021).

10. Kessner, D., Chambers, M., Burke, R., Agus, D. & Mallick, P. ProteoWizard: open source software for rapid proteomics tools development. Bioinformatics 24, 2534–2536 (2008).

11. Pluskal, T., Castillo, S., Villar-Briones, A. & Oresic, M. MZmine 2: modular framework for processing, visualizing, and analyzing mass spectrometry-based molecular profile data. BMC Bioinformatics 11, 395 (2010).

12. Schmid, R. et al. Ion Identity Molecular Networking in the GNPS Environment. bioRxiv 2020.05.11.088948 (2020) doi:10.1101/2020.05.11.088948.

13. Nothias, L.-F. et al. Feature-based molecular networking in the GNPS analysis environment. Nat. Methods 17, 905–908 (2020).

14. Wang, M. et al. Sharing and community curation of mass spectrometry data with Global Natural Products Social Molecular Networking. Nat. Biotechnol. 34, 828–837 (2016).

15. Bolyen, E. et al. Reproducible, interactive, scalable and extensible microbiome data science using QIIME 2. Nat. Biotechnol. 37, 852–857 (2019).

16. Martino, C. et al. A Novel Sparse Compositional Technique Reveals Microbial Perturbations. mSystems 4, (2019).

17. van der Hooft, J. J. J., Wandy, J., Barrett, M. P., Burgess, K. E. V. & Rogers, S. Topic modeling for untargeted substructure exploration in metabolomics. Proc. Natl. Acad. Sci. U. S. A. 113, 13738–13743 (2016).

18. Wandy, J. et al. Ms2lda.org: web-based topic modelling for substructure discovery in mass spectrometry. Bioinformatics 34, 317–318 (2018).

19. Rogers, S. et al. Deciphering complex metabolite mixtures by unsupervised and supervised substructure discovery and semi-automated annotation from MS/MS spectra. Faraday Discuss. 218, 284–302 (2019).

20. da Silva, R. R. et al. Propagating annotations of molecular networks using in silico fragmentation. PLoS Comput. Biol. 14, e1006089 (2018).

21. Dührkop, K. et al. SIRIUS 4: a rapid tool for turning tandem mass spectra into metabolite structure information. Nat. Methods 16, 299–302 (2019).

22. Ernst, M. et al. MolNetEnhancer: Enhanced Molecular Networks by Integrating Metabolome Mining and Annotation Tools. Metabolites 9, (2019).

23. Dührkop, K. et al. Systematic classification of unknown metabolites using high-resolution fragmentation mass spectra. Nat. Biotechnol. 39, 462–471 (2021).

24. Djoumbou Feunang, Y. et al. ClassyFire: automated chemical classification with a comprehensive, computable taxonomy. J. Cheminform. 8, 61 (2016).

25. Rohart, F., Gautier, B., Singh, A. & Lê Cao, K.-A. mixOmics: An R package for ‘omics feature selection and multiple data integration. PLoS Comput. Biol. 13, e1005752 (2017).

26. Benjamini, Y. & Hochberg, Y. Controlling the false discovery rate: A practical and powerful approach to multiple testing. J. R. Stat. Soc. 57, 289–300 (1995).

27. Bodein, A., Chapleur, O., Droit, A. & Lê Cao, K.-A. A Generic Multivariate Framework for the Integration of Microbiome Longitudinal Studies With Other Data Types. Front. Genet. 10, 963 (2019).

28. Petras, D. et al. Chemical Proportionality within Molecular Networks. Anal. Chem. 93, 12833–12839 (2021).

29. Sumner, L. W. et al. Proposed minimum reporting standards for chemical analysis Chemical Analysis Working Group (CAWG) Metabolomics Standards Initiative (MSI). Metabolomics 3, 211–221 (2007).

30. Haid, M. et al. Long-Term Stability of Human Plasma Metabolites during Storage at -80 °C. J. Proteome Res. 17, 203–211 (2018).

31. Goodman, K. et al. Assessment of the effects of repeated freeze thawing and extended bench top processing of plasma samples using untargeted metabolomics. Metabolomics 17, 31 (2021).

32. Wagner-Golbs, A. et al. Effects of Long-Term Storage at -80 °C on the Human Plasma Metabolome. Metabolites 9, (2019).

33. Li, K., Naviaux, J. C., Monk, J. M., Wang, L. & Naviaux, R. K. Improved Dried Blood Spot-Based Metabolomics: A Targeted, Broad-Spectrum, Single-Injection Method. Metabolites 10, (2020).

34. Wu, G., Fang, Y.-Z., Yang, S., Lupton, J. R. & Turner, N. D. Glutathione metabolism and its implications for health. J. Nutr. 134, 489–492 (2004).

35. Haynes, C. A. & De Jesús, V. R. The stability of hexacosanoyl lysophosphatidylcholine in dried-blood spot quality control materials for X-linked adrenoleukodystrophy newborn screening. Clin. Biochem. 48, 8–10 (2015).

